# Quantitative Trait Loci on Chromosome 21 have Pleiotropic Effects on %FEV_1_ and Allergen Polysensitization; asthma related traits in the EGEA study

**DOI:** 10.1101/2020.04.17.20069369

**Authors:** Ayse Ulgen, Christopher Amos

## Abstract

To investigate whether the 21q21 region may contain a quantitative trait locus (QTL) with pleiotropic effect on % predicted FEV1 (forced expiratory volume in 1 second) and SPTQ (number of positive skin test responses to 11 allergens), in 295 EGEA families ascertained through asthmatic probands, we conducted a bivariate linkage analysis using two approaches:

(1) a bivariate variance components (VC) analysis and (2) A combined principal components (CPC) analysis, with 13 microsatellites covering the whole chromosome 21. To identify the genetic variants associated with these traits, we performed family-based association analysis (FBAT) for the second principal component (PC2) using two microsatellites and 27 SNPs belonging to three candidate genes, located in the vicinity of the linkage peak. Univariate linkage analyses showed suggestive evidence of linkage to %FEV1 and SPTQ at two positions. Bivariate VC linkage analysis of these phenotypes led to an increase in linkage signals as compared to univariate analysis at the same positions. The peaks obtained by the CPC led to similar results as the full bivariate VC approach; evidence for linkage mainly coming from PC2. The strongest association signal, using single marker analysis for PC2, was obtained with D21S1252 (p=0.003 for global test; p=0.004 for allele 2 and p=0.001 for allele 11) and rs2834213 of IFNGR2 (p=0.003), these two loci being 3 Mb apart. Multi-marker analysis further strengthened this finding. These results indicate that at least two genetic factors may be involved in SPTQ and %FEV1 variability but further genotyping is needed to better understand these findings.

## INTRODUCTION

According to the World Health Organization, more than 339 million people are living with asthma globally and the numbers are increasing, especially in the Western countries (Douwes and Pearce, 2014; Douwes et al., 2011). Asthma is one of the major non-communicable diseases and it is the most common chronic disease of childhood, affecting 14% children (WHO; www.who.int, Fong MK et al, 2020). Asthma is a multifactorial and complex disease associated with intermediate phenotypes involved in immune response to inflammation and lung function. Although the genetic component of asthma and asthma-related phenotypes has long been established [Vercelli 2008], the extent to which the genetic factors involved are common or specific to these phenotypes is unclear. To date, multiple genetic factors have been identified, with more than 100 loci linked to asthma and asthma-related phenotypes of potential interest in genome-wide-association studies (GWAS) [Hernandez-Pacheco N et al., 2019; Kabesh et al., 2020; Zhu Z et al, 2021]. One interesting feature of these published genome screens is that a given chromosomal region is often linked to various asthma-associated phenotypes across studies, suggesting that these phenotypes may share genetic determinants. However, the formal characterization of pleiotropic effects of genes underlying two or more asthma-related phenotypes has received little attention [Ziegler, et al, 2001; Bouzigon, et al. 2007; Ferreira, et al. 2006; Aschard H, et al. 2009; Dixon and Poytner, 2016].

A previous genome-wide scan conducted in 295 French families from the Epidemiological study on the Genetic and Environmental factors of Asthma (EGEA) for asthma and seven asthma-associated phenotypes detected a linkage signal at the same marker position in the 21q21 region for two asthma-related phenotypes: forced expiratory volume in 1 second percent predicted (%FEV_1_) and a measure of polysensitization to allergens (SPTQ), suggesting that these traits may share genetic determinants [Bouzigon, et al. 2004].

Multivariate genetic linkage analyses and genome-wide association analyses of correlated phenotypes have been shown to improve power of detecting genes with small effects where these genes may be missed with univariate analyses [Allison, et al. 1998; Marlow, et al. 2003; Kochunov P et al, 2010; Wu T et al, 2010; Kochunov P et al, 2011; Yang Q and Wang Y 2012; Deliu M et al, 2016; Salidas et al, 2018; Fatumo S et al, 2019]. Several methods have been proposed to conduct bivariate linkage analysis, among which are the bivariate variance component (VC) approach [Amos, et al. 2001; Bauman, et al. 2005] and a combined analysis of principal components (PCs) [Mangin, et al. 1998]. Using a model with fixed genetic effects in a single phase known cross, [Mangin, et al. 1998] showed the asymptotic equivalences between the combined principal component approach and the bivariate test for linkage. However, these methods were not compared analytically in a VC model. A previous limited simulation study showed similar power for VC and CPC methods [Gorlova, et al. 2002]. We have further studied the asymptotic distribution of the bivariate VC test statistic under the null hypothesis of no linkage and defined upper and lower bounds of this statistic, since the asymptotic distribution of this test is more complex in VC model than in linear model.

In the present study, we tested for pleiotropic genetic determinants that may influence both %FEV_1_ and SPTQ on chromosome 21q21 in the French EGEA families using the bivariate VC analysis and combined analysis of principal components. This linkage analysis was followed by an association analysis with candidate genes in the linkage region.

## METHODS

### FAMILY SAMPLE

The protocol of the EGEA data collection has been described elsewhere [Kauffmann, et al. 2001; Kauffmann, et al. 1997]. The sample examined by the present study consisted of 291 nuclear families ascertained trough at least one asthmatic subject. The inclusion criteria for asthma have been described in details elsewhere [Kauffmann, et al. 1997]. Subjects answered a detailed questionnaire regarding respiratory symptoms and treatment based on international standardized questionnaires. Biological and physiological tests were performed on each participant. Written informed consent was obtained from all subjects participating to the study under an Institutional Review Board-approved protocol.

### PHENOTYPES ANALYZED

Skin-prick tests were performed for 11 allergens (including moulds, indoors and outdoors allergens). A positive response was defined as a wheal size exceeding that of the negative control by ≥ 3 mm [Maccario, et al. 2003] A quantitative score (SPTQ) was constructed by counting the number of positive responses to allergens and thus measuring the degree of polysensitization. SPTQ originally included 12 classes (from 0 to 11) but classes 4 to 11 were combined into the last category because of small sample size. Prior to the analysis, SPTQ was adjusted for relevant covariates including age and sex using multiple regression. These regression models, including main effects and interaction terms, were built separately in three groups (parents, children offspring < 16 years of age, offspring ≥16 years of age) as discussed in Bouzigon et al. [Bouzigon, et al. 2004].

Spirometric measures were carried out for adults and children separately. A survey for adults were conducted according to the European Respiratory Health Survey protocol [Burney, et al. 1994] and [Quanjer 1983] and a survey for children were conducted according to Polgar and Weng [Polgar and Weng 1979]The best of three pre-bronchodilator FEV_1_ measures was used to calculate a percentage of predicted FEV_1_ values (%FEV_1_) based on age, height and gender [Polgar and Weng 1979; Quanjer 1983].

Since, %FEV_1_ and SPTQ showed departures from normality with significant kurtosis for both traits (p < 6.10^−7^), a probit transformation was applied to each phenotype prior to linkage analysis to normalize their distribution [Peng, et al. 2007].

### GENOTYPING

Genotyping of chromosome 21 was done at CNG (Centre National de Génotypage at Evry), with five microsatellites from the original scan and eight additional markers for fine-mapping. The markers were distributed at an average distance of 3 cM and had an average heterozygosity of 75%. After rigorous genotype quality control, the final sample for the present analysis included 291 families (1301 subjects) with at least one asthmatic subject, comprising 566 genotyped parents (97.3% of all parents) and 718 genotyped sibs. In addition to the microsatellite data, three candidate genes genotyped for a total of 27 SNPs and spanning a 33 - 42 Mb region around the 43 cM linkage peak of 21q21 were available in the EGEA study (genotyping done at CNG with the TaqMan® SNP technology). These genes included: 4 SNPs in interferon (alpha and beta) receptor 2 (*IFNAR2*), 15 SNPs in the interleukin 10 receptor, beta (*IL10RB*) and 10 SNPs in the interferon gamma receptor 2 (*IFNGR2*) (see Supplementary Table S1). Additionally, supplementary table S2 shows the microsatellites and SNPs also belonging to the *ADAMTS1, ADAMTS5* and *ICOSL* genes.

## STATISTICAL ANALYSES

### LINKAGE ANALYSIS

Genetic and environmental components of variance for each trait and genetic and environment correlations between the two traits were estimated by variance decomposition using maximum likelihood methods implemented in the multipoint quantitative trait linkage software package (ACT) (http://www.epigenetic.org/Linkage/act.html) [de Andrade 1998].

We first conducted univariate linkage analysis of %FEV_1_ and SPTQ (adjusted and probit transformed values) using the VC method. We then carried out bivariate linkage analysis using bivariate VC and combined analysis of principal components (CPC) of the two phenotypes.

The univariate VC method separates the total variation of a trait into genetic and environmental components and evaluates linkage by comparing a model incorporating both a genetic additive variance 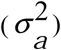 at a putative QTL linked to marker(s) and a polygenic 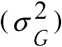 component with a purely polygenic model (QTL variance, 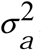, being set to zero) by a likelihood ratio test (LRT). Minus twice the natural logarithm of this likelihood ratio follows asymptotically a one-sided chi-square with one degree of freedom. This chi-square divided by 2ln10 is a LOD score.

The univariate and bivariate variance component models are described in details in Amos *et al* [Amos, et al. 2001] and Williams *et al* [Williams and Blangero 1999]. For the univariate model, under the null hypothesis, the parameters estimated are the mean, μ, the additive genetic variance, 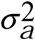, the polygenic variance 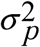, and the environmental variance 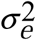. For the bivariate test, under the null hypothesis, the marker linked to the QTL parameters are restricted to be equal to zero 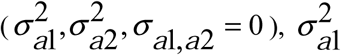 and 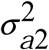 being the additive genetic variance related respectively to trait1 and trait2 and *σ*_*a*1,*a*2_ the additive genetic covariance between these traits. Under the alternative hypothesis, the three parameters are estimated with the constraints: 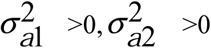, and 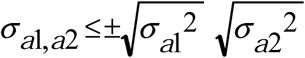. The test for genetic linkage is constructed by a LRT. The distribution of the bivariate linkage test statistic depends whether a constraint is imposed on the genetic correlation between the traits. When the correlation is unconstrained, the asymptotic distribution of the bivariate test statistic, under the null hypothesis that the linked-genetic components and covariance are zero, is the supremum of a 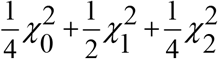 process (See Appendix). However, computing the probability distribution function of this supremum, in order to get quantiles or P-values, is a difficult task, so we used a mixture of 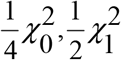, and 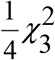 as an approximation of the bivariate test statistic distribution. This approximation was shown, by simulations, to be an upper limit of the distribution [Amos, et al. 2001]. When the genetic correlation between the traits is constrained to be zero, or any other value, the distribution of the bivariate linkage test statistic follows, under the null hypothesis, a mixture of 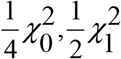, and 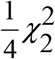 and we define this as the lower bound of the test statistic distribution (See Appendix).

The idea of considering PC analysis of phenotypes prior to linkage analysis to obtain a set of uncorrelated PCs and obtaining a combined test statistic was proposed by Mangin et al [Mangin, et al. 1998]. These authors showed that the likelihood-ratio test used to test for the presence of a pleiotropic QTL modelled as a fixed genetic effect for a given mating type is asymptotically equivalent to the sum of likelihood-ratio tests of univariate analyses applied to the principal components of the phenotypes. Here, we applied this method in a VC model and called this approach as the CPC method. In a first step, a principal components (PC) analysis was applied to %FEV_1_ and SPTQ, to obtain two uncorrelated principal components. These components were then subjected to independent univariate linkage analyses based on the VC method. In a second step, a combined test statistic, CPC test, was constructed by summing the univariate VC likelihood-ratio (LR) test statistics obtained for each of the two 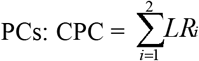, where the distribution of each LR_i_ test, under the null hypothesis of no linkage, is a mixture of 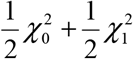. Since the two principal components are independent, the asymptotic distribution of the combined PC test, CPC, is a mixture of 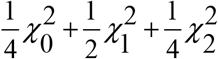.

### FAMILY-BASED ASSOCIATION TEST

Association between quantitative phenotypes (%FEV_1_, SPTQ, PC1 and PC2) and genetic polymorphisms (microsatellites and SNPs) was assessed using the family-based association test (FBAT) method [Lange, et al. 2002]. This method was applied to test for association in the presence of linkage. We tested each marker under additive, dominant and recessive models and the best fitting model was selected. Following single marker analysis, we carried out multi-marker analyses using FBAT-LC [Xu, et al. 2006]. This method allows testing multiple markers simultaneously without haplotype reconstruction. In brief, the FBAT-LC method proposed by Xu et al, is based on a linear combination of single-marker FBAT test statistics using data-driven weights, where marker weight derivation is based on the conditional mean model [Lange, et al. 2003].

## RESULTS

### DESCRIPTIVE STATISTICS

The phenotypic characteristics of 718 genotyped siblings belonging to 291 families are shown in Table I. Among these siblings, fifty-three percent were males and their mean age was 16.0 ± 7.7 (SD) years. The proportion of asthmatic siblings was 54.2%. The proportion of siblings having at least one positive skin prick test was 67.4%. The mean of %FEV_1_ was 96.6 (SD=13.3) and SPTQ was 2.6 (SD=1.4) before adjustment on age and sex. The total number of sibs with phenotypic information was 694 for %FEV_1_, 705 for SPTQ, and 681 when both phenotypes were jointly considered.

**Table I.**
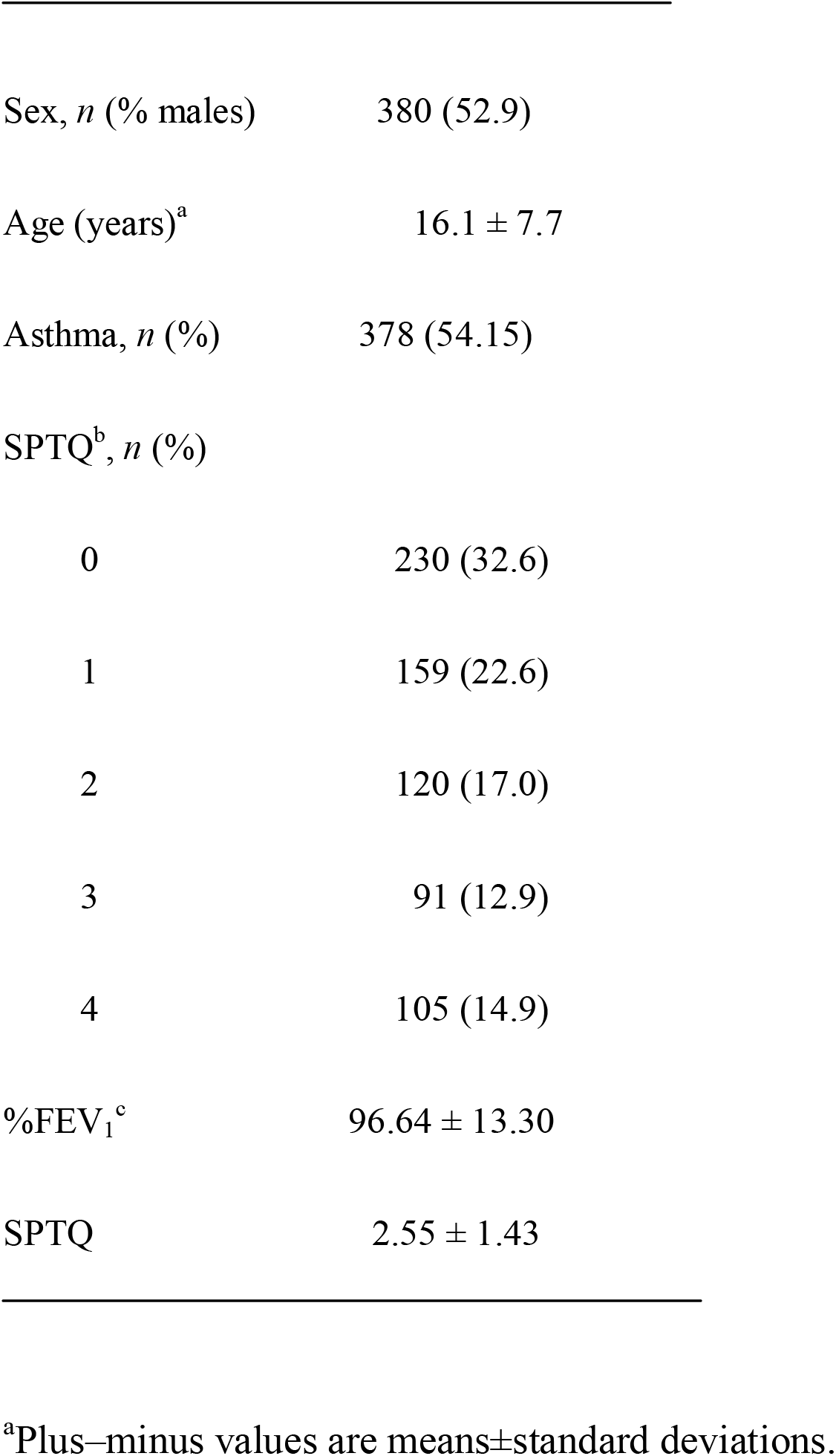
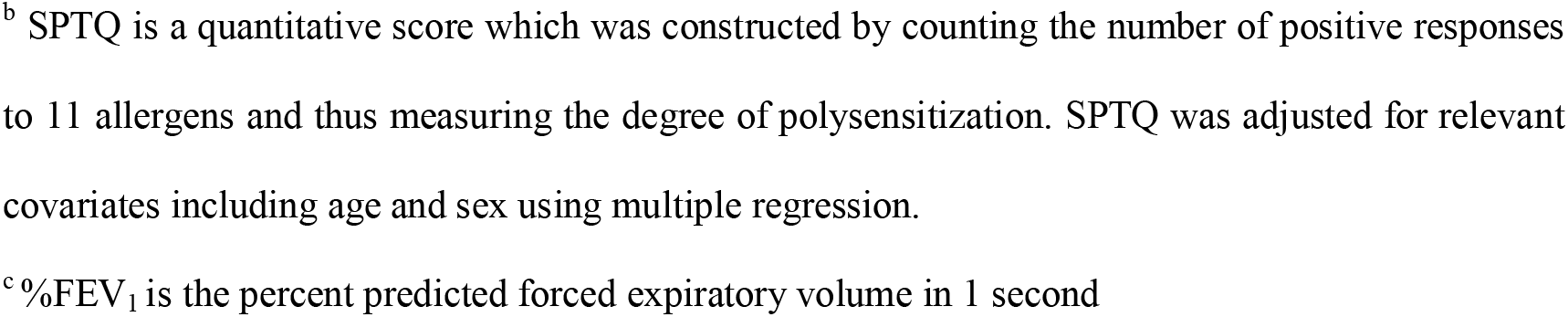
**Phenotypic characteristics of 718 genotyped siblings in 291 families ascertained with ≥ 1 asthmatic subject**

### LINKAGE ANALYSIS

Heritability estimates, (h2), were significant for both probit transformed and age, sex adjusted phenotypes (P < 1.0×10^−4^) and were equal to 38% (SD=3.3) for SPTQ and 57% (SD=3.2) for %FEV_1_. The genetic correlations between %FEV_1_ and SPTQ was positive (ρG = 0.2), whereas the environmental correlation was negative (ρE = -0.2).

Univariate and bivariate linkage analysis results for SPTQ and %FEV_1_ are shown in Figure 1 and Table II. Univariate VC linkage analyses showed evidence for linkage to %FEV_1_ at two positions: D21S265 at 25.5 cM (LOD = 2.2, p = 0.0008) and D21S1252 at 43 cM (LOD=1.9, p=0.002). Univariate VC linkage analysis of SPTQ showed lower evidence for linkage than for %FEV_1_ but at two positions adjacent to the two latter ones; D21S1914 at 24.4 cM (LOD = 1.07, p = 0.01) and D21S1895 at 41.1 cM (LOD = 1.10, p = 0.01).

**Figure 1.**
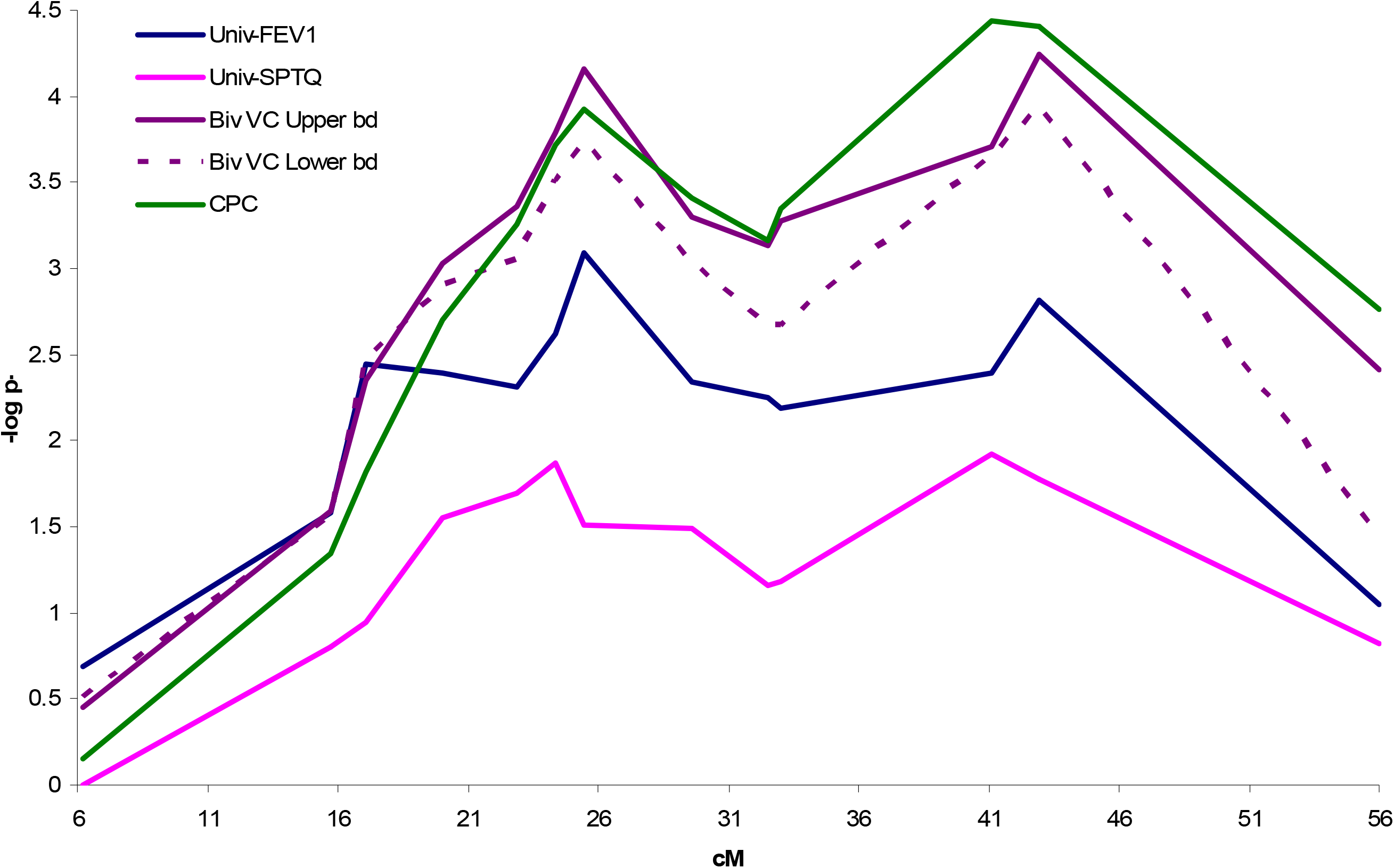
Variance component results of chromosome 21 linkage analysis comparing univariate %FEV_1_, univariate SPTQ, bivariate VC upper bound ; bivariate VC lower bound and CPC analysis, conducted in 291 EGEA families with at least one asthmatic subject. –Log10 p-values are shown on the vertical axis and map distances (in cM) on the horizontal axis.

**Table II.** **P-values for univariate and bivariate linkage analyses conducted for %FEV_1_^*^, SPTQ^+^, PC1^@^ PC2 and CPC, VC (Upper bound), VC (Lower bound) in 291 French EGEA families**

| Marker | Position<br>(Decode cM) | Univariate |  |  |  | Bivariate |  |  |
| --- | --- | --- | --- | --- | --- | --- | --- | --- |
|  |  | %FEV<sub>1</sub><sup>υ</sup> | SPTQ<sup>υ</sup> | PC1<sup>υ</sup> | PC2<sup>υ</sup> | CPC<sup>ψ</sup> | VC(Upper bd)<sup>ξ</sup> | VC (Lower bd)<sup>ψ</sup> |
| D21S1911 | 6.2 | 0.20 | - | 0.45 | 0.49 | 0.70 | 0.35 | 0.30 |
| D21S1899 | 15.7 | 0.03 | 0.16 | 0.30 | 0.02 | 0.05 | 0.03 | 0.03 |
| D21S1905 | 17.1 | 0.004 | 0.11 | 0.14 | 0.01 | 0.02 | 0.004 | 0.003 |
| D21S1922 | 20.0 | 0.004 | 0.03 | 0.15 | 0.001 | 0.002 | 9.0×10<sup>-4</sup> | 0.001 |
| D21S1257 | 22.9 | 0.005 | 0.02 | 0.15 | 3.0×10<sup>-4</sup> | 5.0×10<sup>-4</sup> | 4.0×10<sup>-4</sup> | 9.0×10<sup>-4</sup> |
| D21S1914 | 24.4 | 0.002 | **0.01** | 0.10 | 1.0×10<sup>-4</sup> | 2.0×10<sup>-4</sup> | 2.0×10<sup>-4</sup> | 3.0×10<sup>-4</sup> |
| D21S265 | 25.5 | **8.0×10<sup>-4</sup>** | 0.03 | 0.12 | **7.0×10<sup>-5</sup>** | **1.0×10<sup>-4</sup>** | **7.0×10<sup>-5</sup>** | **2.0×10<sup>-4</sup>** |
| D21S1258 | 29.6 | 0.005 | 0.03 | 0.14 | 2.0×10<sup>-4</sup> | 4.0×10<sup>-4</sup> | 5.0×10<sup>-4</sup> | 9.0×10<sup>-4</sup> |
| D21S263 | 32.5 | 0.006 | 0.07 | 0.27 | 2.5×10<sup>-4</sup> | 7.0×10<sup>-4</sup> | 7.0×10<sup>-4</sup> | 0.002 |
| D21S1909 | 33.0 | 0.006 | 0.07 | 0.30 | 1.5×10<sup>-4</sup> | 4.5×10<sup>-4</sup> | 5.0×10<sup>-4</sup> | 0.002 |
| D21S1895 | 41.1 | 0.004 | **0.01** | 0.17 | **2.0×10<sup>-5</sup>** | **4.0×10<sup>-5</sup>** | 2.0×10<sup>-4</sup> | 2.0×10<sup>-4</sup> |
| D21S1252 | 43.0 | 0.002 | 0.02 | 0.12 | **2.0×10<sup>-5</sup>** | **4.0×10<sup>-5</sup>** | **6.0×10<sup>-5</sup>** | **1.0×10<sup>-4</sup>** |
| D21S266 | 56.0 | 0.09 | 0.15 | - | 5.0×10<sup>-4</sup> | 0.002 | 0.004 | 0.04 |

\* %FEV<sub>1</sub> is the percent predicted forced expiratory volume in 1 second
<sup>+</sup> SPTQ is a quantitative score which was constructed by counting the number of positive responses to 11 allergens and thus measuring the degree of polysensitization. SPTQ was adjusted for relevant covariates including age and sex using multiple regression.
<sup>@</sup> PC1 and PC2 are the two principal components obtained from %FEV<sub>1</sub> and SPTQ
<sup>υ</sup>P-value for the univariate models are calculated as 50:50 mixture of $\chi^2$ distribution (0,1) df.
<sup>ξ</sup>P-value for the bivariate upper bound is calculated as 0.25:0.50:0.25 mixture of $\chi^2$ distribution (0,1,3) df.
<sup>ψ</sup>P-value for the bivariate lower bound is calculated as 0.25:0.50:0.25 mixture of $\chi^2$ distribution (0,1,2) df.

Bivariate VC linkage analysis of %FEV_1_ and SPTQ showed an increase in LOD scores as compared to univariate analyses with highest LOD scores obtained at the two previous positions (Figure 1): D21S265 (25.5 cM) with LOD scores ranging between 3.3 (p = 2.0×10-4) and 4.2 (p = 7.0 ×10^−5^) for the lower and upper bound of test statistics respectively and D21S1252 (43.0 cM) with LOD scores ranging between 3.48 (p = 0.0001) and 4.3 (p = 6.0×10^−5^). Parameter estimates at D21S265 and D21S1252 linked QTL are presented in Table III. Using the unconstrained model, the QTL covariance (*σ* _*a*1, *a*2_) for each of the two markers was positive (0.52-0.55) whereas the polygenic covariance (*σ* _*p*1, *p*2_) was of opposite sign, varying from -0.85 to -0.98. The proportion of total variance explained by the QTL varied from 33% to 36% for %FEV_1_ and from 23% to 29% for SPTQ.

**Table III.** Bivariate Variance Components Parameter Estimates for D21S265 and D21S1252 markers.

| Hypothesis | | | $\sigma^2_{P1}$ | $\sigma^2_{P2}$ | $\sigma_{P1,P2} (\rho_{p1,p2})$ | $\sigma^2_{a1}$ | $\sigma^2_{a2}$ | $\sigma_{a1,a2} (\rho_{a1,a2})$ | $\sigma^2_{E1}$ | $\sigma^2_{E2}$ | lnL | LRT | P-value<sup>ξ</sup> |
| --- | --- | --- | --- | --- | --- | --- | --- | --- | --- | --- | --- | --- | --- |
| Markers Position (cM) |  |  | %FEV<sub>1</sub> | SPTQ |  | %FEV<sub>1</sub> | SPTQ |  | %FEV<sub>1</sub> | SPTQ |  |  |  |
| H0<sup>ψ</sup> |  |  | 0.57 | 0.38 | -0.01 (-0.03) | 0.00 | 0.00 | 0.00 | 0.41 | 0.63 | -650.10 |  |  |
| D21S265 | 25.5 | H1 (Upper bd) | 0.23 | 0.13 | -0.17 (-0.98) | 0.36 | 0.23 | 0.15 (0.52) | 0.40 | 0.64 | -640.52 | 19.16 | 7.0×10<sup>-5</sup> |
|  |  | H1 (Lower bd) | 0.25 | 0.15 | -0.03 (-0.16) | 0.35 | 0.22 | 0.00 (0.00) | 0.40 | 0.64 | -642.55 | 15.10 | 2.0×10<sup>-4</sup> |
| D21S1252 | 43.0 | H1 (Upper bd.) | 0.22 | 0.09 | -0.14 (-0.85) | 0.35 | 0.29 | 0.17 (0.55) | 0.42 | 0.63 | -640.33 | 19.54 | 6.0×10<sup>-5</sup> |
|  |  | H1 (Lower bd) | 0.27 | 0.07 | -0.03 (-0.22) | 0.33 | 0.29 | 0.00 (0.00) | 0.39 | 0.65 | -642.09 | 16.02 | 1.0×10<sup>-4</sup> |

Where $\sigma^2_p$ = polygenic variance, $\sigma_{P1,P2}$ = polygenic covariance, $\rho_{p1,p2}$ = polygenic correlation, $\sigma^2_a$ = additive variance, $\sigma_{a1,a2}$ = additive covariance, $\rho_{a1,a2}$ = QTL correlation, $\sigma^2_E$ = environmental variance
Mean value for %FEV<sub>1</sub> and SPTQ under H0; $\mu$ (H0) = -0.008 (%FEV<sub>1</sub>) and 0.013 (SPTQ)
<sup>ξ</sup> P-value for the bivariate upper bound is calculated as 0.25:0.50:0.25 mixture of $\chi^2$ distribution (0,1,3) df.
P-value for the bivariate lower bound is calculated as 0.25:0.50:0.25 mixture of $\chi^2$ distribution (0,1,2) df.

The principal components (PC) analysis of %FEV_1_ and SPTQ showed that the first and second PCs contributed respectively to 51% and 49% of the overall phenotypic variance. On the first PC (PC1), the coefficients for the two phenotypes were of opposite sign, (0.71 for %FEV_1_ and –0.71 for SPTQ), while on PC2, the coefficients were positive. CPC analysis showed highest evidence for linkage at the same locations as the bivariate method, at 25.5 cM at D21S265 (p = 1.0×10-4) and at 43 cM for D21S1252 (p = 3.9×10^−5^) and at 41.1 cM position at D21S1895 (p = 3.6×10^−5^).

The bivariate VC and CPC methods led to similar results but with a slightly higher evidence for pleiotropic QTLs at 25.6 cM with bivariate VC and at 41.1 and 43 cM with CPC. As can be seen in Table II, for the combined CPC, evidence for linkage came almost solely from PC2 at (p=7.0×10-5 at 25.5 cM and p=2.0×10-5 at 41.1 and 43 cMs) while no significant lod score for PC1 was observed. This led us to further study PC2 for association analysis.

### ASSOCIATION ANALYSIS

The list of 27 SNPs belonging to the three candidate genes spanned a 0.185 Mb region from 33.55 to 33.73 Mb (Supplementary Table S1) between the two linkage peaks. None of these SNPs showed departure from Hardy-Weinberg equilibrium.

Univariate association of PC2 with the microsatellite markers located at the highest linkage peaks and the 10 SNPs of the candidate genes showed association signals with D21S1252 (p=0.003 with a global test under a dominant model) and two SNPs within *IFNGR2* (p=0.02 with rs9976971 and p=0.06 with rs2284553 under an additive model) (See Table IV and Supplementary Table S2 for all markers). When examining each allele separately at D21S1252, allele 2 was positively associated with PC2 (p=0.004) while allele 11 was negatively associated (p=0.001). Interestingly, this microsatellite is located within the *CLDN14* gene.

**Table IV.**
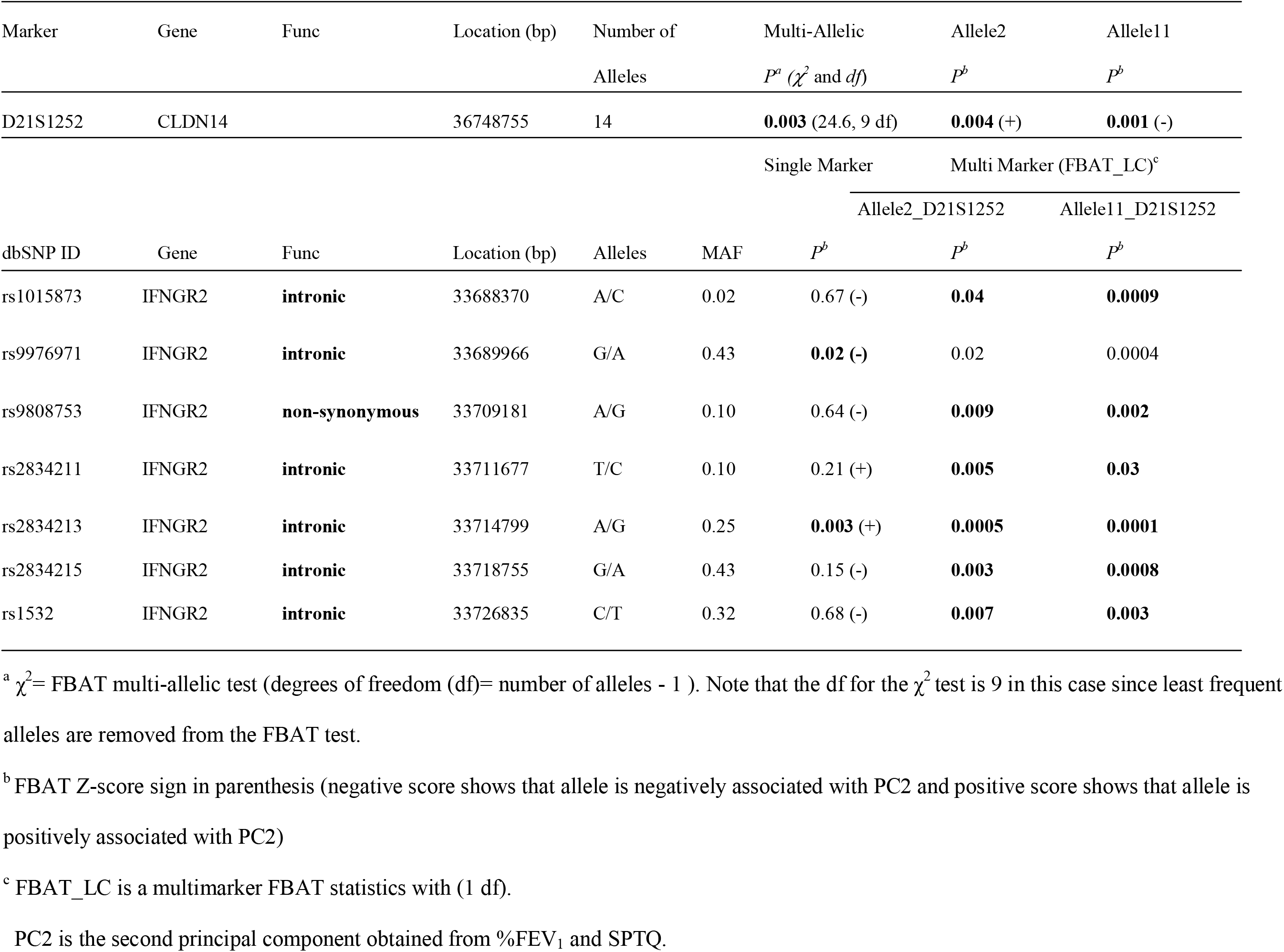
**Results of single and multi marker family-based association (FBAT) analysis for PC2 with the microsatellite marker D21S1252 and SNPs of the *IFNGR2* gene on 21q21, for the best-fitting model.**

This single-marker association analysis was extended to two marker analysis using FBAT_LC by examining jointly each of the SNPs belonging to *IFNGR2* with either allele 2 or 11 from D21S1252. Higher evidence of association was consistently observed for each of *IFNGR2* SNPs in combination with allele 11 as compared with allele 2 of D21S1252. The strongest signal was obtained when considering jointly allele 11 of D21S1252 and *IFNGR2*_rs2834213 (p=0.0001).

Examination of the LD pattern between D21S1252 alleles and *IFNGR2* SNPs showed no LD (LD coefficient r^2^ being < 0.2). The 10 SNPs within *IFNGR2* showed also low pairwise LD except for three pairs of SNPs (see Table V and Figure 2).

**Table V.**
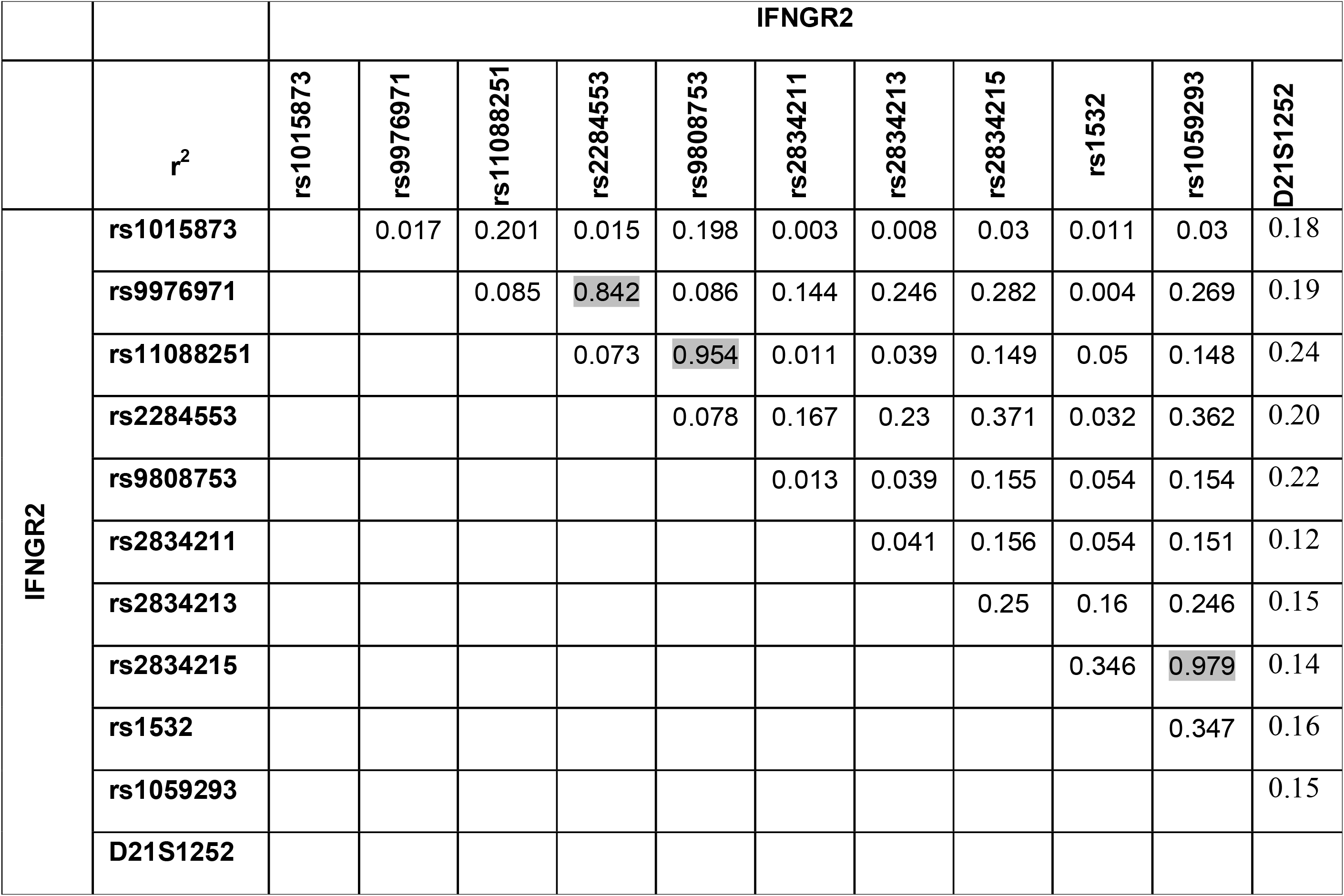
Linkage disequilibrium (r^2^) between *IFNGR2* SNPs.

**Figure 2.**
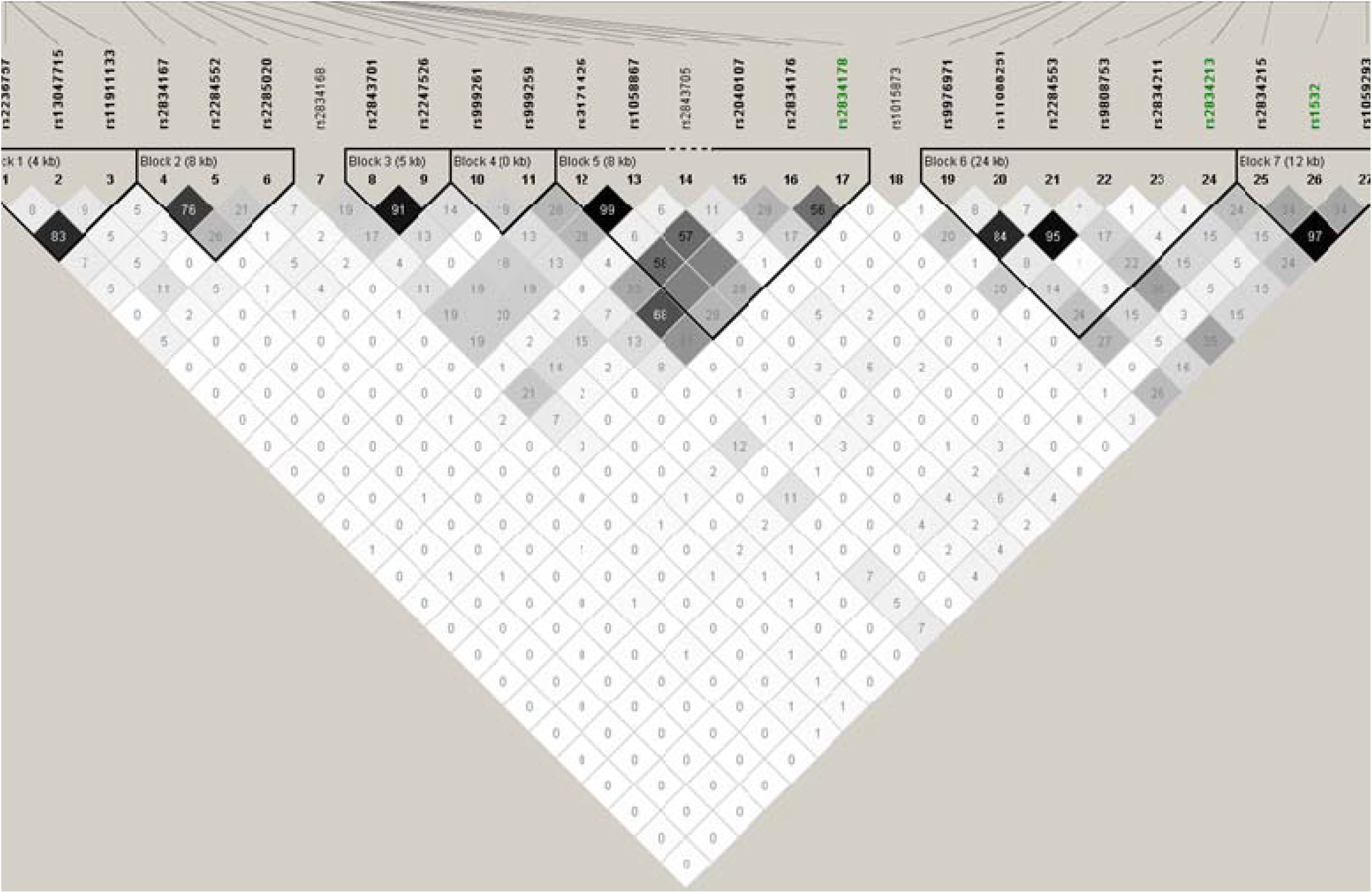
LD measured by r^2^ for *IFNAR2, IL10RB* and *IFNGR2* genes.

## DISCUSSION

The present study is the first one that examined allergy polysensitization and lung function in a bivariate fashion. Bivariate linkage analysis revealed two potential pleiotropic QTLs at 25 cM and 43 cM. Further, association between a combination of lung function values and measures of sensitization was formed with genetic variants at two loci, D21S1252 and *IFNGR2*, 3 Mb distant from each other, close to the 43 cM linkage peak. To our knowledge, this study is also the first attempt to test association between *IFNGR2* gene and %FEV_1_ and allergen polysensitization. These results indicate that two genes in the 21q21 region influence lung function and allergen polysensitization.

Bivariate linkage analysis of %FEV_1_ and SPTQ was carried out using two different methods: a bivariate variance component approach and a combined principal component analysis. Both bivariate linkage analyses led to an increase in linkage signals of the same magnitude at the two positions previously detected by univariate analyses. Simulation studies have shown that VC multivariate approaches are more powerful than univariate analyses when the traits are not very highly positively correlated, this power being highest when correlation relative to QTL and polygenic components underlying two phenotypes are of opposite sign [Amos, et al. 2001; Gorlova, et al. 2002]. Thus, bivariate analysis of %FEV_1_ and SPTQ, which show small phenotypic correlation, may be particularly useful to detect genes with pleiotropic effects on these phenotypes. One disadvantage of the VC approach is that it may be difficult to apply to more than two phenotypes since the number of VC parameters to be estimated increases geometrically as the number of phenotypes increases which can create convergence problems when maximizing the likelihood. Therefore, a potential advantage of the CPC approach is that it can be easily applied to more than two phenotypes although covariance and variances cannot be estimated. Moreover, this approach can be used with other test statistics [Bouzigon, et al. 2007; Aschard, et al, 2009].

Both bivariate VC and CPC approaches assume multivariate normality of the phenotypes and departure from normality may lead to inflation of type I error [Allison, et al. 1999]. However, probit transformation of each phenotype was used here to ensure the validity of the normality assumption.

Replication of linkage results across studies are important in supporting the actual involvement of linkage regions. We have considered all previously reported linkage peaks with P ≤ 0.01 by published genome scans performed to date in more than 20 different populations. Five different genome screens have reported linkage signals to 21q. At 21q21, around the first linkage peak for lung function, atopic asthma and specific skin prick tests (Postma et al, 2005, Pillai et al, 2006 and Blumenthal et al 2006). The 21q22 region around our second linkage peak was also reported linked to asthma and several atopy related phenotypes (specific IgE, specific skin prick tests) in three different populations (CSGA, Hutterites and German families) [The Collaborative Study on the Genetics of Asthma, 1997; Blumenthal, et al. 2006; Ober, et al. 1999; Wjst 1999]. This is in agreement with our results.

The QTL detected on 21q22, *IFNGR2* gene, encodes the non-ligand-binding beta chain of the gamma interferon (*IFNG*) receptor. *IFNG* has been implicated to be associated to several asthma related phenotypes and its role on immune and allergic responses has been established [Malerba and Pignatti 2005; Teixeria LK et al, 2005]. Human interferon-gamma receptor is a heterodimer of *IFNGR1* and *IFNGR2*. [Nakao, et al. 2001] investigated the association of a coding variant, *Gln64Arg* of the *IFNGR2* gene, with atopic asthma in Japanese child population but did not find any significant associations whereas [Gao, et al. 1999] found an association of *Gln64Arg* variant with total serum IgE levels in the British population. Interestingly, the *Gln64Arg* is also represented in our analysis sample as rs9808753 but no association signal was obtained for PC2 with this SNP. Most recently, Daley et al [Daley, et al. 2009] reported an association between *IFNGR2* gene and atopic asthma using four asthma population samples from Canada and Australia. Finally, data from the International HapMap consortium suggest that, 6 out 10 snps of *IFNGR2* gene in the EGEA data are within a large block of linkage disequilibrium, capturing most of the variation in the *IFNGR2* gene.

D21S1252 microsatellite is located within one of the markers belonging to the Claudin 14 (*CLDN14*) gene. [Tsukita and Furuse 2002] reported that primary and secondary dysfunction of the claudin-based barrier in various organs can cause various pathological conditions. The facilitation of antigen presentation via a paracellular pathway is hypothesised to trigger or aggravate chronic inflammation. In asthma it has been proposed that various peptidases contained in allergens, such as house dust mites and fungus, cleave the extracellular loops of claudins/occludin to affect the TJ barrier [Wan, et al. 1999], [Robinson, et al. 2001]. Along the same lines, it is possible that dysfunction of the claudin-based barrier in the epidermis is involved in the pathogenesis of epidermal inflammation such as atopic dermatitis [Leung 1995]. Recently, Rodriguez et al [Rodriguez, et al. 2009] carried out a meta analysis to show strong associations of fillagrin(*FLG*) mutations with eczema and asthma. This may be another example of a disrupted barrier which may support the hypothesis with the *CLDN14* gene.

In conclusion, the present study shows that both use of quantitative phenotypes of %FEV_1_ and SPTQ, and using two different multivariate approaches, indicated support for a pleiotropic effect at two different positions on chromosome 21. An association analysis further indicated a moderate association signal for PC2 with two loci: D21S1252 and rs2834213 belonging to *IFNGR2*. Multi-marker analysis with these two loci strengthened this signal, especially when considering jointly allele 11 of D21S1252 and rs2834213. Although the p values obtained are generally low, these are considered nominal uncorrected p values and hence these results need to be followed up with a confirmatory study and these results bode well for a replication study. Future functional analysis and further genotyping may help to identify the causal variants in the *IFNGR2* gene for %FEV_1_ and allergen polysensitization.

## Data Availability

Data and quality management: Inserm ex-U155 (Egea1): J Hochez; Inserm U 780, Villejuif: N Le Moual, C Ravault; Inserm U 794: N Chateigner; Grenoble: J Ferran.

## EGEA cooperative group

### Coordination

F Kauffmann; F Demenais (genetics); I Pin (clinical aspects).

### Respiratory epidemiology

Inserm U 700, Paris M Korobaeff (Egea1), F Neukirch (Egea1); Inserm U 707, Paris: I Annesi-Maesano; Inserm U 780, Villejuif: F Kauffmann, N Le Moual, R Nadif, MP Oryszczyn; Inserm U 823, Grenoble: V Siroux.

### Genetics

Inserm U 393, Paris: J Feingold; Inserm U 535, Villejuif: MH Dizier; Inserm U 794, Evry: E Bouzigon, F Demenais; CNG, Evry: I Gut, M Lathrop.

### Clinical centers

Grenoble: I Pin, C Pison; Lyon: D Ecochard (Egea1), F Gormand, Y Pacheco; Marseille: D Charpin (Egea1), D Vervloet; Montpellier: J Bousquet; Paris Cochin: A Lockhart (Egea1), R Matran (now in Lille); Paris Necker: E Paty, P Scheinmann; Paris-Trousseau: A Grimfeld, J Just.

### Data and quality management

Inserm ex-U155 (Egea1): J Hochez; Inserm U 780, Villejuif: N Le Moual, C Ravault; Inserm U 794: N Chateigner; Grenoble: J Ferran.

This study was funded by INSERM, the French Ministry of Higher Education and Research, University of Evry, Fondation pour la Recherche Médicale (ACE20061209064) and the French National Agency for Research (ANR 05-SEST-020-02/05-9-97) and ANR 06-CEBS-029-2).

## Appendix

### 1. Asymptotic Distribution of the bivariate VC test

The bivariate variance component model leads to a non-regular likelihood with a nuisance parameter present only under the alternative. Indeed, the correlation parameter of the variance-covariance matrix of the linked QTL disappears when variances are equal to 0. This type of non-regular likelihood was studied by Davies (1977, Biometrika 64: 247-254; 1987, Biometrika 74: 33-43). However, since the likelihood function is regular when the correlation *ρ* is known, the asymptotic law of the LRT for the bivariate VC model can be studied as the supremum of the asymptotic law of the likelihood ratio test given *ρ*.

Let *l*_*N*_ (*θ, τ, ρ*) denote the log likelihood function of the bivariate VC model where *N* is the number of independent families, *ρ* is the QTL additive genetic correlation, 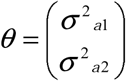 and*τ* are the other unknown parameters. Assuming that the true value of *τ* does not lie on the boundary of the parameter space and that *ρ* is known, we can restrict our attention to *θ*. Following Self and Liang (1987), let *U*_*N*_ (*θ* ; *ρ*) and −*I* _*N*_ (*θ* ; *ρ*) denote the first and second derivatives of the log likelihood function given. In the neighborhood of the null hypothesis*θ*_0_ = (0,0)′, the likelihood ratio test given *ρ* is asymptotically equal to 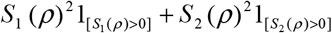 Where

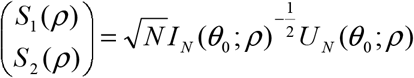

And we found an asymptotic probability function that is a mixture 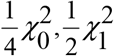, and 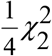. However, when the bivariate variance component model is unconstraint, the correlation parameter is estimated under the alternative hypothesis so we get

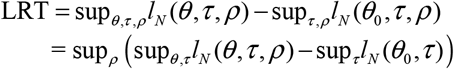

Therefore, the LRT of the bivariate variance component model is asymptotically equal to 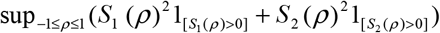 which is the supremum of a 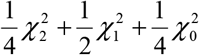 process.

For the upper bound of the distribution the results obtained by simulation in Amos et al 2001 is used. Their simulations study showed that the 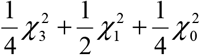 quantiles are conservative for the unconstraint bivariate VC test, therefore 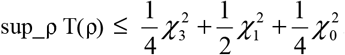.

## Figure legends

**Supplementary Table S1 :** Microsatellite markers and SNPs belonging to the *IFNAR2, IL10RB, IFNGR2* genes.

| Marker | Gene | position (Mb) | MAF | HW_P |
| --- | --- | --- | --- | --- |
| D21S1911 |  | 15.062606 |  |  |
| D21S1899 |  | 18.989572 |  |  |
| D21S1905 |  | 19.866424 |  |  |
| D21S1922 |  | 21.220615 |  |  |
| D21S1257 |  | 23.735675 |  |  |
| D21S1914 |  | 24.544187 |  |  |
| D21S265 |  | 25.841347 |  |  |
| D21S1258 |  | 27.741551 |  |  |
| D21S263 |  | 31.143785 |  |  |
| D21S1909 |  | 31.455116 |  |  |
| rs2236757 | IFNAR2 | 33.546786 | 0.287 | 0.110 |
| rs13047715 | IFNAR2 | 33.547459 | 0.174 | 0.700 |
| rs11911133 | IFNAR2, IL10R | 33.551044 | 0.321 | 0.124 |
| rs2834167 | IL10RB | 33.562657 | 0.277 | 0.018 |
| rs2284552 | IFNAR2, IL10R | 33.565951 | 0.23 | 0.190 |
| rs2285020 | IL10RB | 33.570992 | 0.416 | 0.239 |
| rs2834168 | IL10RB | 33.57266 | 0.319 | 0.975 |
| rs2843701 | IL10RB | 33.574326 | 0.448 | 0.279 |
| rs2247526 | IL10RB | 33.579556 | 0.458 | 0.136 |
| rs999261 | IL10RB | 33.588029 | 0.175 | 0.865 |
| rs999259 | IL10RB | 33.588361 | 0.485 | 0.247 |
| rs3171425 | IL10RB | 33.590616 | 0.412 | 0.227 |
| rs1058867 | IL10RB | 33.59125 | 0.41 | 0.187 |
| rs2843705 | IL10RB | 33.596424 | 0.044 | 0.246 |
| rs2040107 | IL10RB | 33.597831 | 0.291 | 0.772 |
| rs2834176 | IL10RB | 33.598239 | 0.422 | 0.153 |
| rs2834178 | IL10RB | 33.59926 | 0.295 | 0.240 |
| rs1015873 | IFNGR2 | 33.68837 | 0.023 | 0.033 |
| rs9976971 | IFNGR2 | 33.689966 | 0.427 | 0.896 |
| rs11088251 | IFNGR2 | 33.692679 | 0.101 | 0.586 |
| rs2284553 | IFNGR2 | 33.698564 | 0.403 | 0.640 |
| rs9808753 | IFNGR2 | 33.709181 | 0.104 | 0.791 |
| rs2834211 | IFNGR2 | 33.711677 | 0.107 | 0.860 |
| rs2834213 | IFNGR2 | 33.714799 | 0.254 | 0.329 |
| rs2834215 | IFNGR2 | 33.718755 | 0.426 | 0.516 |
| rs1532 | IFNGR2 | 33.726835 | 0.319 | 0.465 |
| rs1059293 | IFNGR2 | 33.731562 | 0.427 | 0.427 |
| D21S1895 |  | 35.272854 |  |  |
| D21S1252 |  | 36.748728 |  |  |
| D21S266 |  | 41.606426 |  |  |

1 reference: dbSNP UCSC hg18 (stsMap) NCBI Build 36.1 or Estimation from contig position (CNG files), HW P: Hardy-Weinberg P-value and MAF: Minor Allele Frequency

**Supplementary Table S2 :** P-values for microsatellite markers and SNPs belonging to the *IFNAR2, IL10RB, IFNGR2* genes, for the best-fitting model.

| Marker | Gene | position (Mb) | MAF | HW_P | P-val |
| --- | --- | --- | --- | --- | --- |
| D21S1911 |  | 15.062606 |  |  | 0.2573 |
| D21S1899 |  | 18.989572 |  |  | 0.3583 |
| D21S1905 |  | 19.866424 |  |  | 0.1987 |
| D21S1922 |  | 21.220615 |  |  | 0.7330 |
| D21S1257 |  | 23.735675 |  |  | 0.4662 |
| D21S1914 |  | 24.544187 |  |  | 0.1040 |
| D21S265 |  | 25.841347 |  |  | 0.2422 |
| rs229042 | ADAMTS1 | 27.132527 | 0.251 | 0.050 | 0.5489 |
| rs162506 | ADAMTS5 | 27.248811 | 0.335 | 0.928 | 0.8743 |
| D21S1258 |  | 27.741551 |  |  | 0.2536 |
| D21S263 |  | 31.143785 |  |  | 0.3050 |
| D21S1909 |  | 31.455116 |  |  | 0.3048 |
| rs2236757 | IFNAR2 | 33.546786 | 0.287 | 0.110 | 0.6192 |
| rs13047715 | IFNAR2 | 33.547459 | 0.174 | 0.700 | 0.4422 |
| rs11911133 | IFNAR2,<br>IL10R | 33.551044 | 0.321 | 0.124 | 0.2287 |
| rs2834167 | IL10RB | 33.562657 | 0.277 | 0.018 | 0.7570 |
| rs2284552 | IFNAR2,<br>IL10R | 33.565951 | 0.23 | 0.190 | 0.5619 |
| rs2285020 | IL10RB | 33.570992 | 0.416 | 0.239 | 0.2141 |
| rs2834168 | IL10RB | 33.57266 | 0.319 | 0.975 | 0.4930 |
| rs2843701 | IL10RB | 33.574326 | 0.448 | 0.279 | 0.7438 |
| rs2247526 | IL10RB | 33.579556 | 0.458 | 0.136 | 0.8639 |
| rs999261 | IL10RB | 33.588029 | 0.175 | 0.865 | 0.1454 |
| rs999259 | IL10RB | 33.588361 | 0.485 | 0.247 | 0.0598 |
| rs3171425 | IL10RB | 33.590616 | 0.412 | 0.227 | 0.2810 |
| rs1058867 | IL10RB | 33.59125 | 0.41 | 0.187 | 0.2909 |
| rs2843705 | IL10RB | 33.596424 | 0.044 | 0.246 | 0.1754 |
| rs2040107 | IL10RB | 33.597831 | 0.291 | 0.772 | 0.2584 |
| rs2834176 | IL10RB | 33.598239 | 0.422 | 0.153 | 0.0745 |
| rs2834178 | IL10RB | 33.59926 | 0.295 | 0.240 | 0.0082 |
| rs1015873 | IFNGR2 | 33.68837 | 0.023 | 0.033 | 0.6777 |
| rs9976971 | IFNGR2 | 33.689966 | 0.427 | 0.896 | 0.0234 |
| rs11088251 | IFNGR2 | 33.692679 | 0.101 | 0.586 | 0.6385 |
| rs2284553 | IFNGR2 | 33.698564 | 0.403 | 0.640 | 0.0596 |
| rs9808753 | IFNGR2 | 33.709181 | 0.104 | 0.791 | 0.6385 |
| rs2834211 | IFNGR2 | 33.711677 | 0.107 | 0.860 | 0.2070 |
| rs2834213 | IFNGR2 | 33.714799 | 0.254 | 0.329 | 0.0032 |
| rs2834215 | IFNGR2 | 33.718755 | 0.426 | 0.516 | 0.1541 |
| rs1532 | IFNGR2 | 33.726835 | 0.319 | 0.465 | 0.6837 |
| rs1059293 | IFNGR2 | 33.731562 | 0.427 | 0.427 | 0.2443 |
| D21S1895 |  | 35.272854 |  |  | 0.5866 |
| D21S1252 |  | 36.748728 |  |  | 0.0034 |
| D21S266 |  | 41.606426 |  |  | 0.4356 |
| rs2838532 | ICOSL | 44.464293 | 0.345 | 0.889 | 0.0703 |
| rs2298565 | ICOSL | 44.467823 | 0.451 | 0.309 | 0.3479 |
| rs3804033 | ICOSL | 44.472915 | 0.213 | 0.046 | 0.1438 |
| rs3746962 | ICOSL | 44.47584 | 0.083 | 0.057 | 0.5828 |
| rs3746963 | ICOSL | 44.475924 | 0.34 | 0.400 | 0.2178 |
| rs3746963 | ICOSL | 44.475924 | 0.34 | 0.400 | 0.2218 |
| rs2070558 | ICOSL | 44.480085 | 0.314 | 0.162 | 0.5443 |
| (C29940613) | ICOSL | 44.480085 | 0.119 | 0.633 | 0.0908 |
| rs2070561 | ICOSL | 44.482397 | 0.315 | 0.191 | 0.5599 |
| rs378299 | ICOSL | 44.48577 | 0.424 | 0.745 | 0.0778 |
| rs2026880 | ICOSL | 44.489757 | 0.41 | 0.407 | 0.5054 |
| rs3737435 | ICOSL | 44.490032 | 0.273 | 0.570 | 0.1500 |
| rs2014457 | ICOSL | 44.493771 | 0.223 | 0.917 | 0.3028 |
| rs7354779 | ICOSL | 44.495197 | 0.257 | 0.559 | 0.7214 |

1 reference: dbSNP UCSC hg18 (stsMap) NCBI Build 36.1 or Estimation from contig position (CNG files) , HW P: Hardy-Weinberg P-value and MAF: Minor Allele
Frequency

## REFERENCES

Aschard A, Bouzigon E, Corda E, Ulgen A, Dizier MH, Gormand F, Lathrop M, Kauffmann F, Demenais F. 2009. Sex-specific effect of IL9 polymorphisms on lung function and polysensitization. Genes Immun 10(6):559–65. doi:10.1038/gene.2009.46.

The Collaborative Study on the Genetics of Asthma (CSGA).1997. A genome-wide search for asthma susceptibility loci in ethnically diverse populations. 1997 Nat Genet 15(4):389–92.

The Collaborative Study on the Genetics of Asthma (CSGA). 2020. Publisher Correction: A genome-wide search for asthma susceptibility loci in ethnically diverse populations. Nat Genet 52, 1433. doi.org/10.1038/s41588-020-0650-1

Allison DB, Neale MC, Zannolli R, Schork NJ, Amos CI, Blangero J. 1999. Testing the robustness of the likelihood-ratio test in a variance-component quantitative-trait loci-mapping procedure. Am J Hum Genet 65(2):531–44.

Allison DB, Thiel B, St Jean P, Elston RC, Infante MC, Schork NJ. 1998. Multiple phenotype modeling in gene-mapping studies of quantitative traits: power advantages. Am J Hum Genet 63(4):1190–201.

Amos C, de Andrade M, Zhu D. 2001. Comparison of multivariate tests for genetic linkage. Hum Hered 51(3):133–44.

Bauman LE, Almasy L, Blangero J, Duggirala R, Sinsheimer JS, Lange K. 2005. Fishing for pleiotropic QTLs in a polygenic sea. Ann Hum Genet 69(Pt 5):590–611.

Blumenthal MN, Langefeld CD, Barnes KC, Ober C, Meyers DA, King RA, Beaty TH, Beck SR, Bleecker ER, Rich SS. 2006. A genome-wide search for quantitative trait loci contributing to variation in seasonal pollen reactivity. J Allergy Clin Immunol 117(1):79–85.

Bouzigon E, Dizier MH, Krahenbuhl C, Lemainque A, Annesi-Maesano I, Betard C, Bousquet J, Charpin D, Gormand F, Guilloud-Bataille M and others. 2004. Clustering patterns of LOD scores for asthma-related phenotypes revealed by a genome-wide screen in 295 French EGEA families. Hum Mol Genet 13(24):3103–13.

Bouzigon E, Ulgen A, Dizier MH, Siroux V, Lathrop M, Kauffmann F, Pin I, Demenais F. 2007. Evidence for a pleiotropic QTL on chromosome 5q13 influencing both time to asthma onset and asthma score in French EGEA families. Hum Genet 121(6):711–9.

Burney PG, Luczynska C, Chinn S, Jarvis D. 1994. The European Community Respiratory Health Survey. Eur Respir J 7(5):954–60.

Daley D, Lemire M, Akhabir L, Chan-Yeung M, He JQ, McDonald T, Sandford A, Stefanowicz D, Tripp B, Zamar D and others. 2009. Analyses of associations with asthma in four asthma population samples from Canada and Australia. Hum Genet 125(4):445–59.

de Andrade M, Kruskal J, Yu L, et al. 1998. ACT- A computer package for analysis of complex traits. Denver: Am Soc Hum Genet.

Deliu M, Sperrin M, Belgrave D, Custovic A. 2016. Identification of Asthma Subtypes Using Clustering Methodologies. Pulm Ther 2:19–41. DOI10.1007/s41030-016-0017-z

Dixon AE, Poynter ME. 2016. Mechanisms of Asthma in Obesity. Pleiotropic Aspects of Obesity Produce Distinct Asthma Phenotypes. Am J Respir Cell Mol Biol; 54(5):601–8. doi:10.1165/rcmb.2016-0017PS

Douwes J and Pearce N. Epidemiology of Respiratory Allergies and Asthma. Handbook of Epidemiology. 2014 : 2263–2319. doi:10.1007/978-0-387-09834-0_50

Douwes J, Brooks C, van Dalen C, Pearce N. 2011. Importance of Allergy in Asthma: An Epidemiologic Perspective. Current Allergy and Asthma Reports 11: 434

Fatumo S, Carstensen T, Nashiru O, Gurdasani D, Sandhu M, Kaleebu P. 2019. Complimentary Methods for Multivariate Genome-Wide Association Study Identify New Susceptibility Genes for Blood Cell Traits. Front. Genet., 10:334| doi.org/10.3389/fgene.2019.00334

Ferreira MA, O’Gorman L, Le Souef P, Burton PR, Toelle BG, Robertson CF, Martin NG, Duffy DL. 2006. Variance components analyses of multiple asthma traits in a large sample of Australian families ascertained through a twin proband. Allergy 61(2):245– 53.

Fong MK, Welte T for the Forum of International Respiratory Societies (FIRS). 2020. World Lung Day: what, why, and where to? Am J Physiol Lung Cell Mol Physiol 319: L527–L533.

Gao PS, Mao XQ, Jouanguy E, Pallier A, Doffinger R, Tanaka Y, Nakashima H, Otsuka T, Roberts MH, Enomoto T and others. 1999. Nonpathogenic common variants of IFNGR1 and IFNGR2 in association with total serum IgE levels. Biochem Biophys Res Commun 263(2):425–9.

Gorlova OY, Amos CI, Zhu DK, Wang W, Turner S, Boerwinkle E. 2002. Power of a simplified multivariate test for genetic linkage. Ann Hum Genet 66(Pt 5-6):407–17.

Hernandez-Pacheco N, Pino-Yanes M, Flores C. 2019. Genomic Predictors of Asthma Phenotypes and Treatment Response. Front Pediatr. 7:6.

Haapakoski R, Karisola P, Fyhrquist N, Savinko T, Lehtimäki S, Wolff H, Lauerma A, Alenius H. 2013. Toll-like receptor activation during cutaneous allergen sensitization blocks development of asthma through IFN-gamma-dependent mechanisms. J Invest Dermatol.133(4):964–72. doi:10.1038/jid.2012.356.

Kauffmann F, Dizier MH, Annesi-Maesano I, Bousquet J, Charpin D, Demenais F, Ecochard D, Feingold J, Gormand F, Grimfeld A and others. 2001. [Epidemiological study of genetic and environmental factors in asthma, bronchial hyperresponsiveness and atopy. Protocol and potential selection bias]. Rev Epidemiol Sante Publique 49(4):343–56.

Kabesh M, Tost J. Recent findings in the genetics and epigenetics of asthma and allergy. 2020. Seminars In Immunopathology 42:43–60.

Kauffmann F, Dizier MH, Pin I, Paty E, Gormand F, Vervloet D, Bousquet J, Neukirch F, Annesi I, Oryszczyn MP and others. 1997. Epidemiological study of the genetics and environment of asthma, bronchial hyperresponsiveness, and atopy: phenotype issues. Am J Respir Crit Care Med 156(4 Pt 2):S123–9.

Kochunov P, Glahn DC, Hong LE, Lancaster J, Curran JE, Johnson MP, Winkler AM, Holcomb HH, Kent Jr. JW, Mitchell B, Kochunov V, Olvera RL, Cole SA, Dyer TD, Moses EK, Goring H, Almasy L, Duggirala R, Blangero J. 2012. P-selectin expression tracks cerebral atrophy in Mexican-Americans. Front. Genet. doi.org/10.3389/fgene.2012.000652

Kochunov P, Glahn D, Lancaster J, Winkler A, Kent JW, Olvera RL, Cole SA, Dyer TD, Almasy L, Duggirala R, Fox PT, Blangero J. Whole brain and regional hyperintense white matter volume and blood pressure: overlap of genetic loci produced by bivariate, whole-genome linkage analyses. 2010. Stroke 41(10):2137–42. doi:10.1161/STROKEAHA.110.590943

Lange C, DeMeo D, Silverman EK, Weiss ST, Laird NM. 2003. Using the noninformative families in family-based association tests: a powerful new testing strategy. Am J Hum Genet 73(4):801–11.

Lange C, DeMeo DL, Laird NM. 2002. Power and design considerations for a general class of family-based association tests: quantitative traits. Am J Hum Genet 71(6):1330–41.

Leung DY. 1995. Atopic dermatitis: the skin as a window into the pathogenesis of chronic allergic diseases. J Allergy Clin Immunol 96(3):302-18; quiz 319.

Maccario J, Oryszczyn MP, Charpin D, Kauffmann F. 2003. Methodologic aspects of the quantification of skin prick test responses: the EGEA study. J Allergy Clin Immunol 111(4):750–6.

Malerba G, Pignatti PF. 2005. A review of asthma genetics: gene expression studies and recent candidates. J Appl Genet 46(1):93–104.

Mangin B, Thoquet P, Grimsley N. 1998. Pleiotropic QTL Analysis. Biometrics 54, 88-99. Marlow AJ, Fisher SE, Francks C, MacPhie IL, Cherny SS, Richardson AJ, Talcott JB, Stein JF, Monaco AP, Cardon LR. 2003. Use of multivariate linkage analysis for dissection of a complex cognitive trait. Am J Hum Genet 72 (3):561–70.

Mathias RA, Freidhoff LR, Blumenthal MN, Meyers DA, Lester L, King R, et al. CSGA (Collaborative Study of the Genetics of Asthma). 2001. Genome-wide linkage analyses of total serum IgE using variance components analysis in asthmatic families. Genet Epidemiol 20(3):340–55.doi:10.1002/gepi.5.

Nadif R, Zerimech F, Bouzigon E, Matran R. 2013. The role of eosinophils and basophils in allergic diseases considering genetic findings. Curr Opin Allergy Clin Immunol. 13(5):507–13. doi:10.1097/ACI.0b013e328364e9c0.

Nakao F, Ihara K, Kusuhara K, Sasaki Y, Kinukawa N, Takabayashi A, Nishima S, Hara T. 2001. Association of IFN-gamma and IFN regulatory factor 1 polymorphisms with childhood atopic asthma. J Allergy Clin Immunol 107(3):499–504.

Nedelkopoulou N, Dhawan A, Xinias I, Gidaris D, Farmaki E. 2020. Interleukin 10: the critical role of a pleiotropic cytokine in food allergy. 48(4):401–408. DOI:10.1016/j.aller.2019.10.003

Ober C, Hoffjan S. 2006. Asthma genetics 2006: the long and winding road to gene discovery. Genes Immun 7(2):95–100.

Ober C, Tsalenko A, Willadsen S, Newman D, Daniel R, Wu X, Andal J, Hoki D, Schneider D, True K and others. 1999. Genome-wide screen for atopy susceptibility alleles in the Hutterites. Clin Exp Allergy 29 Suppl 4:11–5.

Oryszczyn MP, Bouzigon E, Maccario J, Siroux V, Nadif R, Wright A, Kauffmann F. 2007. Interrelationships of quantitative asthma-related phenotypes in the Epidemiological Study on the Genetics and Environment of Asthma, Bronchial Hyperresponsiveness, and Atopy. J Allergy Clin Immunol;119(1):57–63. doi:10.1016/j.jaci.2006.09.026.

Peng B, Yu RK, Dehoff KL, Amos CI. 2007. Normalizing a large number of quantitative traits using empirical normal quantile transformation. BMC Proc 1 Suppl 1:S156.

Polgar G, Weng TR. 1979. The functional development of the respiratory system from the period of gestation to adulthood. Am Rev Respir Dis 120(3):625–95.

Quanjer P. 1983. Working party on ‘standardized lung function testing’. Bull. Eur. Physiopathol. Respir. 19, 7–10.

Robinson C, Baker SF, Garrod DR. 2001. Peptidase allergens, occludin and claudins. Do their interactions facilitate the development of hypersensitivity reactions at mucosal surfaces? Clin Exp Allergy 31(2):186– 92.

Rodriguez E, Baurecht H, Herberich E, Wagenpfeil S, Brown SJ, Cordell HJ, Irvine AD, Weidinger S. 2009. Meta-analysis of filaggrin polymorphisms in eczema and asthma: robust risk factors in atopic disease. J Allergy Clin Immunol 123(6):1361–70 e7.

Salinas YD, Wang Z, DeWan AT. Statistical Analysis of Multiple Phenotypes in Genetic Epidemiologic Studies: From Cross-Phenotype Associations to Pleiotropy. 2018. American Journal of Epidemiology, Volume 187, Issue 4: 855–863, doi.org/10.1093/aje/kwx296

Sleiman PMA, Hakonarson H. 2010. Recent advances in the genetics and genomics of asthma and related traits. Curr Opin Pediatr. 22(3):307–12.doi:10.1097/MOP.0b013e328339553d.

Teixeira LK, Fonseca BPF, Barboza BA, Viola JPB. 2005. The role of interferon-g on immune and allergic responses Mem. Inst. Oswaldo Cruz vol.100 uppl.1. http://dx.doi.org/10.1590/S0074-02762005000900024

Tsukita S, Furuse M. 2002. Claudin-based barrier in simple and stratified cellular sheets. Curr Opin Cell Biol 14(5):531–6.

Vercelli D. 2008. Discovering susceptibility genes for asthma and allergy. Nat Rev Immunol 8(3):169–82.

Wan H, Winton HL, Soeller C, Tovey ER, Gruenert DC, Thompson PJ, Stewart GA, Taylor GW, Garrod DR, Cannell MB and others. 1999. Der p 1 facilitates transepithelial allergen delivery by disruption of tight junctions. J Clin Invest 104(1):123–33.

Williams JT, Blangero J. 1999. Comparison of variance components and sibpair-based approaches to quantitative trait linkage analysis in unselected samples. Genet Epidemiol 16(2):113–34.

Wjst M. 1999. Specific IgE--one gene fits all? German Asthma Genetics Group. Clin Exp Allergy 29 Suppl 4:5–10.

Wu T, Boezen HM, Postma DS, Los H, Postmus PE, Snieder H, Boomsma DI. 2010. Genetic and environmental influences on objective intermediate asthma phenotypes in Dutch twins. European Respiratory Journal 36: 261–268; DOI:10.1183/09031936.00123909

Xu X, Rakovski C, Xu X, Laird N. 2006. An efficient family-based association test using multiple markers. Genet Epidemiol 30(7):620–6.

Yang Q and Wang Y. Advanced Designs and Statistical Methods for Genetic and Genomic Studies of Complex Diseases. 2012. Methods for Analyzing Multivariate Phenotypes in Genetic Association Studies. J of Probability and Statistics. Volume 2012, Article ID 652569: 1–13. https://doi.org/10.1155/2012/652569

Zhu Z, Hasegawa K, Camargo Jr CA, Liang L. 2021. Investigating asthma heterogeneity through shared and distinct genetics: Insights from genome-wide cross-trait analysis. J Allergy Clin Immunol. 147(3):796–807. doi:10.1016/j.jaci.2020.07.004.

Ziegler A, Böddeker IR, Geller F. 2001. A bivariate Haseman-Elston method and application to the analysis of asthma-related phenotypes on chromosome 5q. Genet Epidemiol. 21 Suppl 1:S216–21. doi:10.1002/gepi.2001.21.s1.s216.

